# Rapid loss of activity but not serum concentration in a passive infusion clinical trial of an HIV neutralizing antibody

**DOI:** 10.1101/2025.09.04.25334949

**Authors:** Sharana Mahomed, Nonhlanhla N Mkhize, Leonid A. Serebryannyy, Tandile Hermanus, Prudence Kgagudi, Haajira Kaldine, Bronwen Lambson, Sandeep R. Narpala, Manjula Basappa, Mike Castro, Bob C Lin, Kevin Carlton, Jason T. Sands, Jason G. Gall, Richard A. Koup, Lynn Morris, Quarraisha Abdool Karim, Penny L. Moore, Salim Abdool Karim, Nicole Doria-Rose

## Abstract

Monoclonal antibodies (mAbs) are a major class of drugs for treatment and prevention of disease. In early clinical trials, the pharmacokinetics (pK) of mAbs are usually assessed by measuring mAb concentration in serum. However, it is not a given that the mAbs will retain full functionality over time, emphasizing the need for integrated PK and functional assessments. In a re-analysis of data from the CAPRISA 012B trial, a previously published phase 1 study evaluating mAbs CAP256V2LS and VRC07-523LS in HIV-negative women, we report an unexpected disconnect between serum bNAb concentrations and HIV neutralization activity of CAP256V2LS, with implications for ongoing assessment of passive immunization trials.

## Main text

HIV broadly neutralizing monoclonal antibodies (bNAbs) are being explored as long-acting HIV prevention tools, with multiple early-phase trials underway. The ability of bNAbs to prevent HIV infection has been demonstrated in numerous non-human primate studies, where bNAb levels correlated with protection.^2^ The AMP clinical trials showed that the VRC01 bNAb prevented acquisition of highly sensitive HIV strains.^3^ In published studies of several passively infused bNAbs in clinical trials, serum bNAb concentration and neutralization activity correlated closely over time, as expected.^4-7^

In CAPRISA 012B,^1^ we administered CAP256V2LS alone or with VRC07-523-LS in various regimens, comparing doses, co-administration of two antibodies, and route of delivery. This phase 1 trial was conducted at the CAPRISA eThekwini Clinical Research Site in Durban, South Africa. The protocol was reviewed and approved by the University of KwaZulu-Natal Biomedical Research Ethics Committee and the South African Health Products Regulatory Authority, and registered on the Pan African Clinical Trial Registry, PACTR202003767867253. Serum concentrations of each antibody were measured using anti-idiotype reagents specific to each bNAb, and neutralization titers (observed ID50) against strains that were sensitive to only one or the other antibody were measured with the TZM-bl Env-pseudovirus assay.

As previously reported,^1^ CAP256V2LS serum concentrations remained detectable for up to six months with an estimated half-life of 43 days, and no anti-drug antibody (ADA) was detected. However, comparison of the dynamics of neutralization vs serum concentration in serum showed a more rapid decline of neutralization titer than antibody concentration for CAP256V2LS, but not for VRC07-523LS. This is illustrated in Figure 1 for subjects receiving a 20 mg/kg single dose of each antibody co-administered. Predicted ID50 values were calculated as bNAb serum concentration divided by the IC50 of the tested virus. A faster decline in CAP256V2LS neutralization titers compared to serum concentration was reflected by the difference in predicted and observed ID50, and increased over time, indicative of loss of functional activity of the antibody present in the serum. In contrast, observed VRC07-523LS neutralization activity in the same participants tracked with the serum concentration, with the ratio between titer and concentration remaining consistent across all time points. The discordance between CAP256V2LS serum concentrations and neutralization activity was observed across all study arms including varying product dosages, routes of administration, presence or absence of recombinant human hyaluronidase (rHuPH20, product name Enhanze) used for subcutaneous administration, and single vs. co-bNAb administration (Supplementary Figures S1-S4).

**Figure 1.**
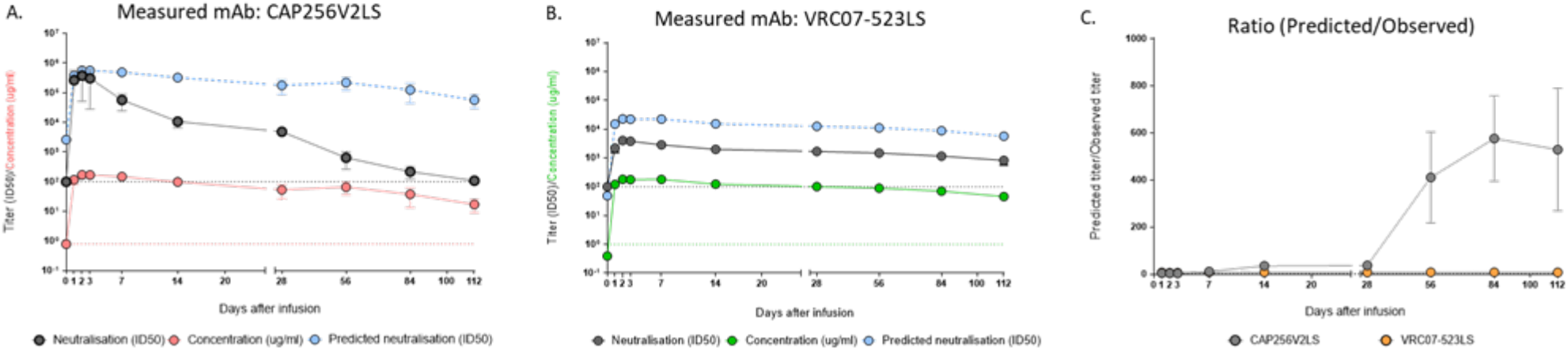
Comparison of bNAb concentration and neutralization activity in CAPRISA 012B participants. All subjects shown received a dual bNAb dose of 20 mg/kg body weight of CAP256V2LS (n=4) and VRC07-523LS (n=4), administered subcutaneously in the presence of Enhanze on Day 0. **A**. Concentration (µg/ml) (red line), measured neutralization titer (ID50) (black line) against HIV-CE2103_E8, and predicted neutralization (ID50) (concentration divided by the IC50, blue line) for CAP256V2LS. **B**. Concentration (µg/ml) (green line), measured neutralization titer (ID50) (black line) against HIV-Q769.D22, and predicted neutralization (ID50) (blue line) for VRC07-523-LS. **C**. Ratio of predicted ID50 to observed ID50 for CAP256V2LS (grey line) and VRC07-523-LS (orange line). Error bars show the standard deviation from the mean of participant data (n=4). The lower limit of detection (LOD) for the neutralisation assay is defined as an ID50 of 100. For PK assays the LOD was 0.8µg/ml and is shown by the red dotted lines

This loss of *in vivo* functionality was specific to CAP256V2LS, even within donors who received both CAP256V2LS and VRC07-523LS. Potential mechanisms include immune complex formation, non-specific binding to serum proteins, loss of post-translational modifications such as tyrosine sulfation, and structural changes such as proteolytic cleavage, despite engineering to resist degradation.^8^ This phenomenon has not been observed in studies of other bNAbs, including VRC07-523LS (Figure 1) as well as 10E8VLS, N6LS, VRC01, and VRC01LS ^4-7^, which maintain proportional neutralization activity relative to serum concentrations. Defining the mechanism for this selective loss in neutralization of CAP256V2LS is the subject of ongoing studies, given that neutralization activity is a critical determinant of bNAb efficacy in preventing HIV^9^. Ongoing results from the CAPRISA 012C trial,^10^ expected in 2025, will clarify whether the observed decline in neutralization activity compromises its overall preventative efficacy. Investigators evaluating other monoclonal antibodies for infectious diseases and other conditions should remain vigilant for similar patterns and incorporate functional assays alongside assessments of serum antibody concentrations to better predict efficacy.

## Data Availability

All data produced in the present study are available upon reasonable request to the authors

## Authors’ Contributions

SM and NM wrote the first draft of the manuscript. PM and NDR acted as senior authors, offering overarching scientific leadership, conceptual guidance, and critical revision of the work. SAK is the Principal Investigator of CAPRISA 012B. QAK and SM are Co-Principal investigators. SM contributed to clinical investigations and sample collection. NNM, PM, NDR, LS, TH, PK, HK, BL, SN, MB, MC, BCL, KC, JS, JG conducted the laboratory assays and contributed to data analysis and interpretation. RAK, KC, PLM, NDR and LM contributed to antibody development. All authors reviewed the final draft of the manuscript and approved the final version.

## Acknowledgments

We thank the study team and the study participants for their contribution to the CAPRISA 012B trial and for furthering HIV prevention research. We thank Halozyme Therapeutics for providing the rHuPH20 (Enhanze), and we thank Mayank Patel and Kenny Israni for their collaborations. We thank the NIAID VRC Vaccine Production Program, Office of Regulatory Science, Clinical Trials Program and Vaccine Immunology Program and the VRC Pilot Plant, operated by the Vaccine Clinical Materials Program (VCMP), Leidos Biomedical Research, Inc. for study product provision. We thank Labcorp for their collaboration on the immunogenicity testing. We thank the CAPRISA 012B protocol and study team, the CAPRISA 012B Protocol Safety Review Team and the CAPRISA 012B Data and Safety Monitoring Board members for providing trial oversight.

## Supplementary Figures

**Figure S1.**
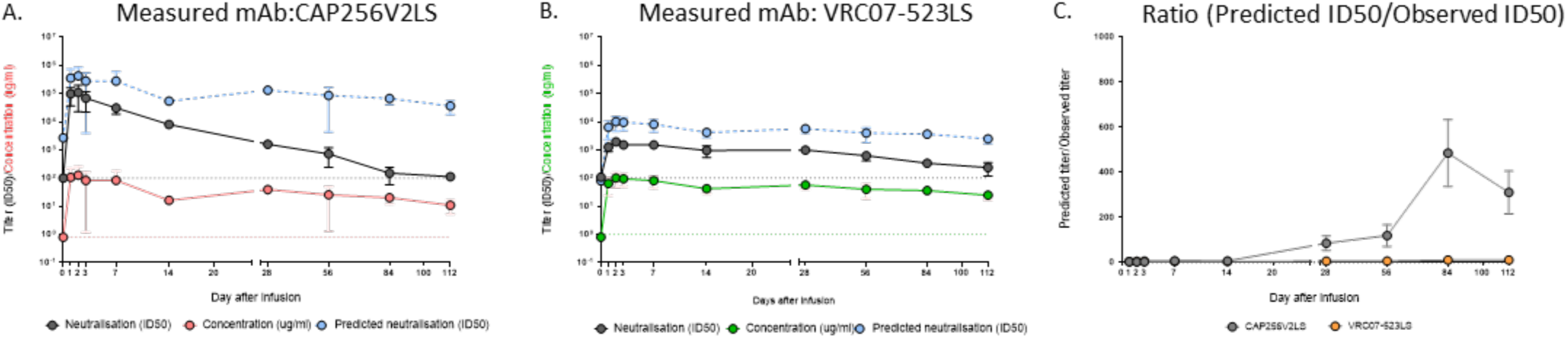
Dual bNAb administration: 10mg/kg SC each, CAP256V2LS and VRC07-523LS. Antibodies were administered together in the presence of Enhanze on Day 0. **A**. Concentration (µg/ml) (red line), measured neutralization titer (ID50) (black line) against HIV-CE2103_E8, and predicted neutralization (ID50) (concentration divided by the IC50, blue line) for CAP256V2LS. **B**. Concentration (µg/ml) (green line), measured neutralization titer (ID50) (black line) against HIV-Q769.D22, and predicted neutralization (ID50) (blue line) for VRC07-523LS. **C**. Ratio of predicted ID50 to observed ID50 for CAP256V2LS (grey line) and VRC07-523LS (orange line). Error bars show the standard deviation from the mean of participant data (n=4). The lower limit of detection (LOD) for the neutralisation assay is defined as an ID50 of 100. For PK assays the LOD was 0.8µg/ml and is shown by the red dotted lines

**Figure S2.**
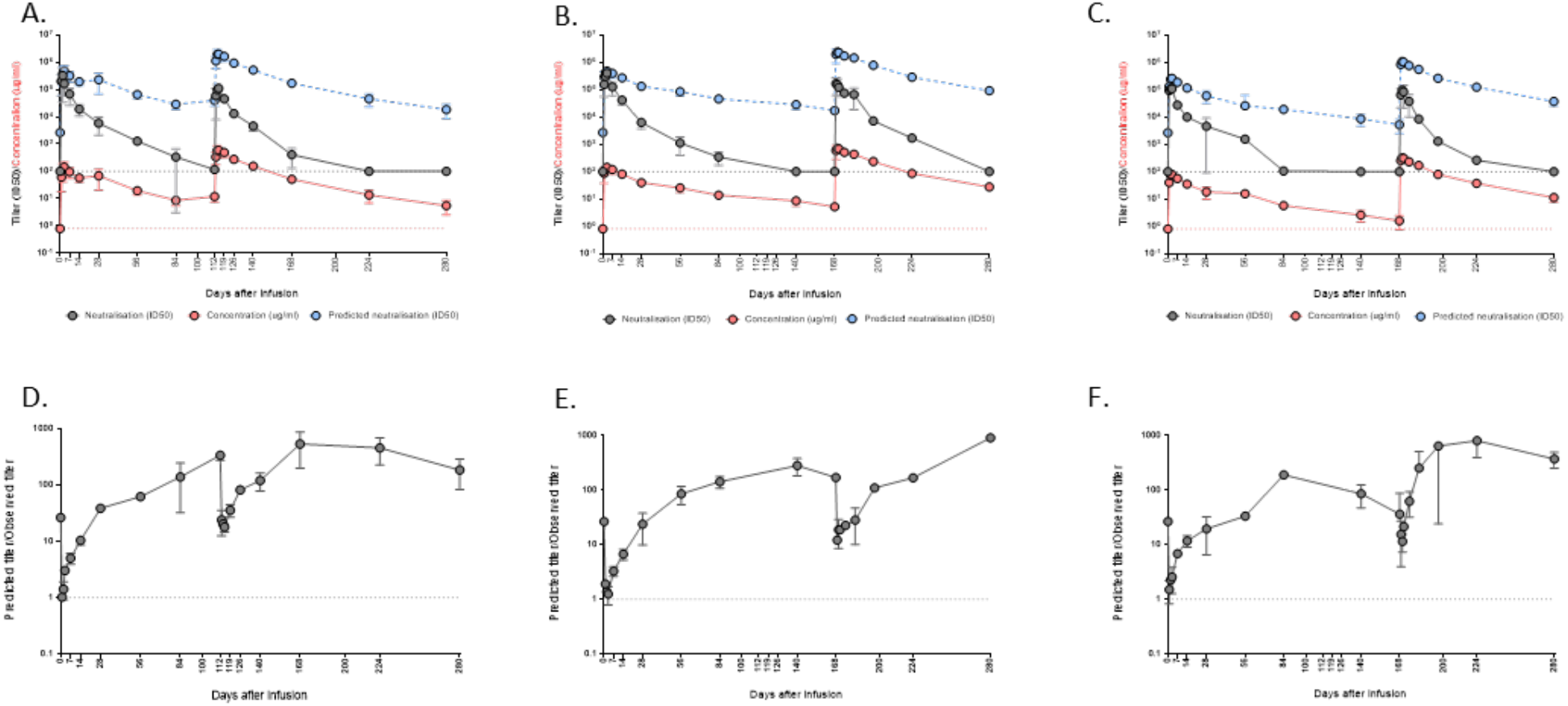
Repeated doses of CAP256V2LS alone at Days 0 and 112 or Day 0 and 168. Participants were given CAP256V2LS subcutaneously, in the presence of recombinant human hyaluronidase (rHuPH20), at doses of **A**. 10mg/kg, with repeated dose at Day 112 or **B**. 10mg/kg, with repeat dose at Day 168 or **C**. 20mg/kg, with repeat dose at Day 168. Red line: concentration (µg/ml). Black line: neutralization titer (ID50) against HIV-CE2103_E8. Blue line: predicted neutralization (ID50). The lower limit of detection (LOD) for the neutralisation assay is defined as an ID50 of 100. For PK assays the LOD was 0.8µg/ml and is shown by the red dotted lines. **D-F**. Ratios of predicted titer (ID50) to observed titer (ID50) for CAP256V2LS, corresponding to above panels. The black dotted line represents ratio (Predicted to Observed)=1. Error bars show the standard deviation from the mean of participant data (n=4).

**Figure S3.**
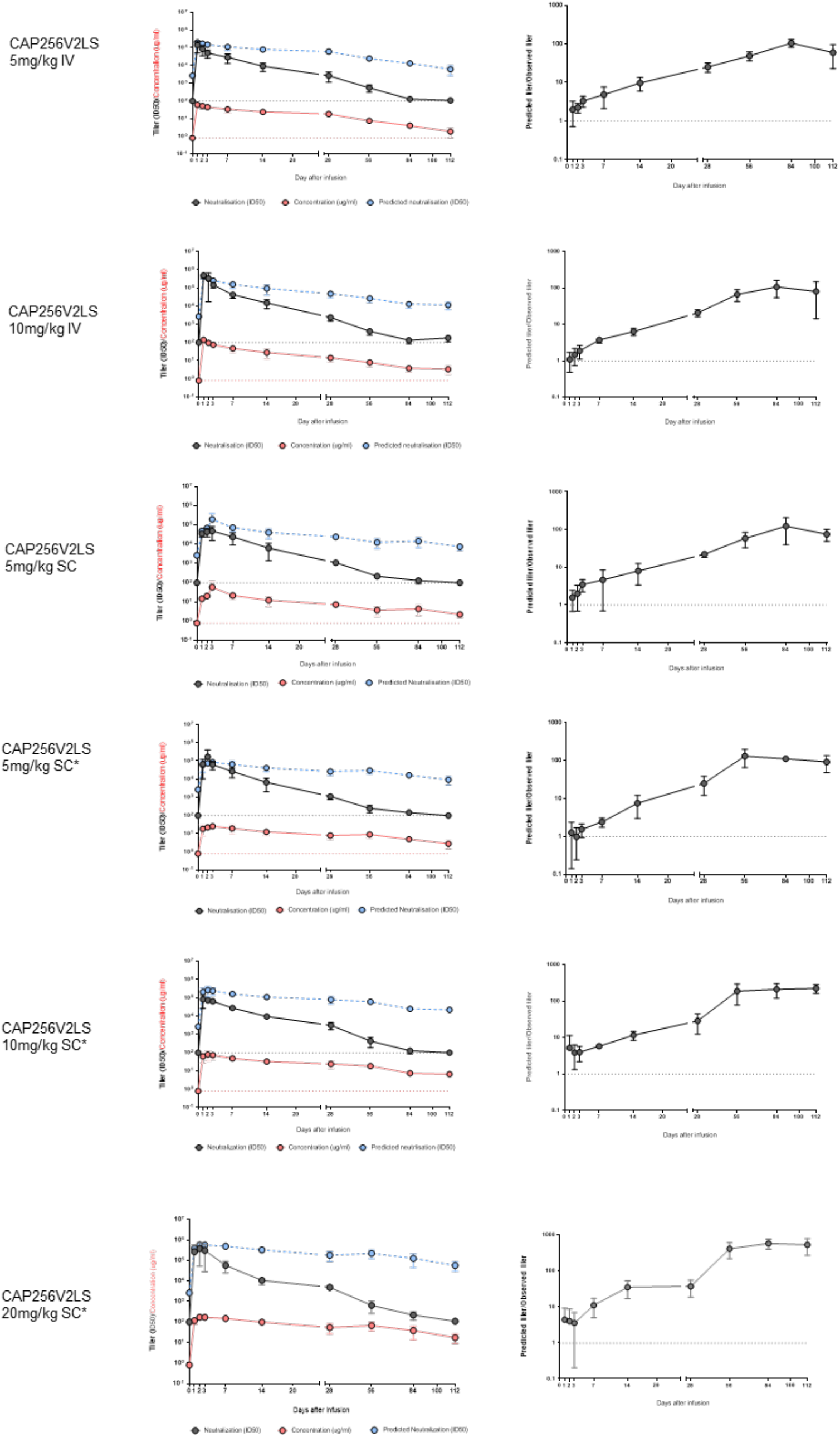
Single doses of CAP256V2LS alone. 4 participants per group received a single dose of CAP256V2LS with the administration route and dosage amount shown. * indicates subcutaneous delivery in the presence of recombinant human hyaluronidase (rHuPH20). Left panels: Red line: concentration (µg/ml). Black line: measured neutralization titer (ID50) against HIV-CE2103_E8. Blue line: predicted neutralization (ID50) (concentration divided by the IC50). The lower limit of detection (LOD) for the neutralisation assay is defined as an ID50 of 100. For PK assays the LOD was 0.8µg/ml and is shown by the red dotted lines. Right panels: Ratios of predicted titer (ID50) to observed titer (ID50) for CAP256V2LS. The black dotted line represents ratio (Predicted to Observed)=1. Error bars show the standard deviation from the mean of participant data (n=4).

**Figure S4.**
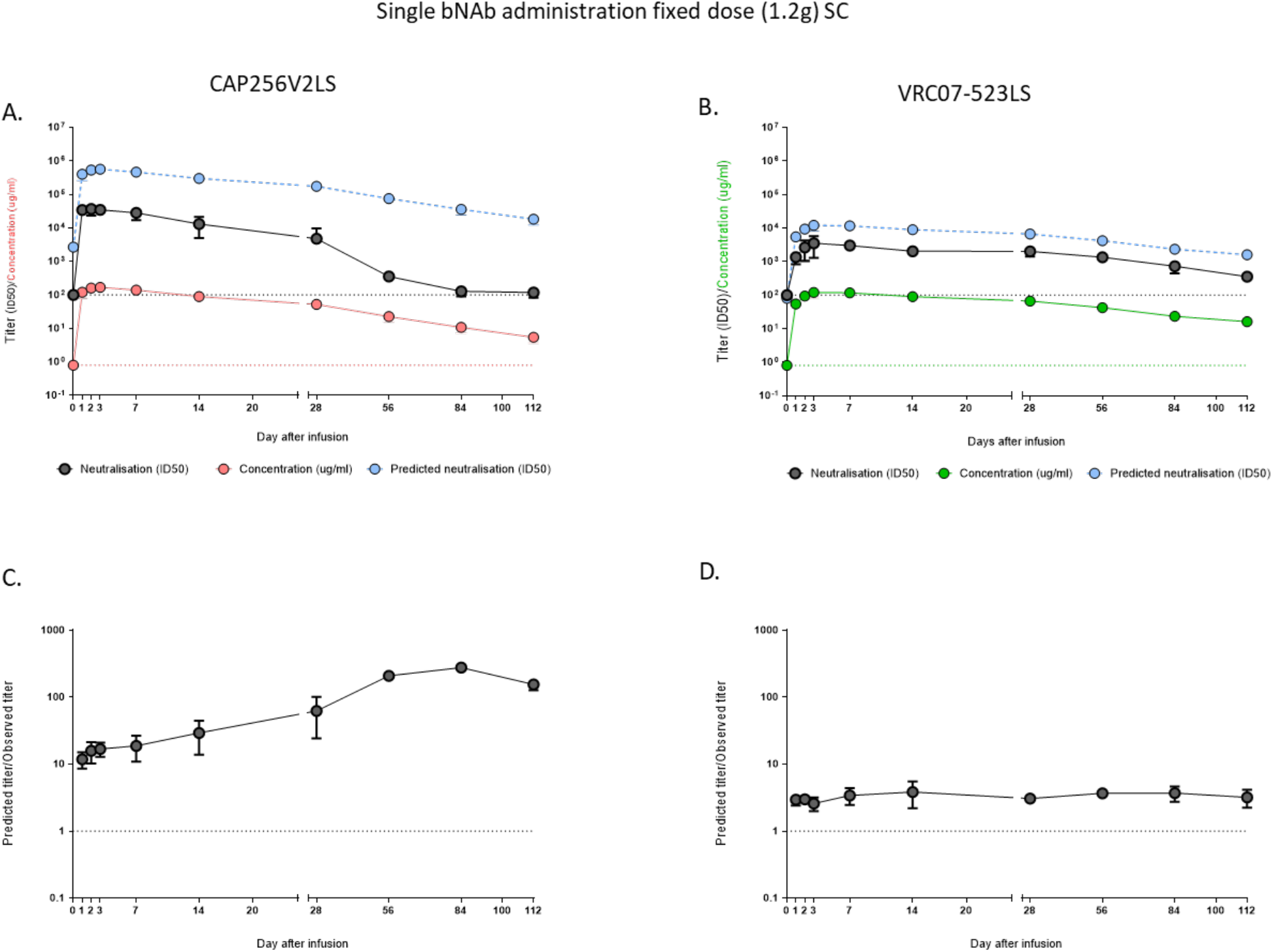
Comparison of concentration and neutralization activity in CAP012B participants after administration of fixed dose CAP256V2LS or VRC07-523LS of 1.2 g. **A**. Lines show concentration (µg/ml) of CAP256V2LS (red line), measured neutralization titer (ID50) against HIV-CE2103 (black line), and predicted neutralization (ID50) (concentration divided by the IC50, blue line). The lower limit of detection (LOD) for the neutralization assay is defined as an ID50 of 100. For PK assays the LOD was 0.8µg/ml (red dotted lines). **B**. Lines show concentration (µg/ml) of VRC07-523-LS (red line), measured neutralization titer (ID50) against HIV-Q769.D422 (black line), and predicted neutralization (ID50) for VRC07-523LS. For PK assay, LOD was 1ug/ml (green dotted line), **C, D**. Ratio of predicted titer (ID50) to observed titer (ID50) for **C**. CAP256V2LS or **D**. VRC07-523-LS. The black dotted line represents ratio (Predicted to Observed) =1. Graphs display neutralization titers and concentrations from four participants per group. Error bars show the standard deviation from the mean of participant data (n=4)

